# Nutritional status and its associated factors among patients with chronic kidney disease attending in a tertiary center of Nepal

**DOI:** 10.1101/2025.08.01.25332706

**Authors:** Susmita Gyawali, Arjun Aryal, Dinesh Koirala

**Author notes:** Correspondence: Susmita Gyawali.

## Abstract

Chronic kidney disease is a global public health concern, characterized by an irreversible deterioration in renal function. Inadequate nutrition and a sedentary lifestyle worsen the vicious cycle of muscle mass and strength loss, which limits the quality of life and rehabilitation opportunities for chronic kidney disease patients. This study aims to assess nutritional status and its association with different factors including physical activity among patients with chronic kidney disease. A cross-sectional study design was used in 290 patients with chronic kidney disease chosen by circular systematic random sampling technique. Standard validated tools: 7-point subjective global assessment was used to assess the nutritional status of the patients using interview and observation techniques. Written informed consent was taken from participants.

Descriptive statistics were used to report socio-demographic variables, chronic kidney disease status-related variables, nutritional status and level of physical activity. The test of association was performed using the chi-square test. The logistic regression model was used to determine factors associated with malnutrition. This study showed, 21.8 to 32% of participants had malnutrition and 16.8% to 26.1% of the participants had low activity level. Male participants were less likely to have malnutrition compared to female participants with chronic kidney disease (AOR 0.451, p-value 0.005). Participants having low activity level were about seven times more likely to have malnutrition compared to those having high activity level (AOR 7.022, p-value <0.001). Similarly, participants having moderate activity level were about three times more likely to have malnutrition compared to those having high activity level (AOR 3.263, p-value 0.005). This study provides evidence of a high burden of malnutrition and explained the relationship between malnutrition and low physical activity among chronic kidney disease patients.

## Introduction

Chronic kidney disease (CKD) is a growing worldwide public health problem and is on the rapid rise due to an increase in diabetes and hypertension. It is characterized by an irreversible decline in renal function that could result in end-stage renal disease, which requires treatment with renal replacement therapy, such as hemodialysis or a kidney transplant. CKD is defined by a Glomerular Filtration Rate (GFR) of < 60 ml/min/1.73m2 and the presence of kidney damage, regardless of the cause, during a period of three or more months [1]

Globally, 1 out of 10 adult people have chronic kidney disease which is responsible for decreasing quality of life, particularly in the later stages of the disease [2]. According to the Global Burden of Disease study, CKD was the 11th leading cause of death in 2019, up from 19th in 1990 accounting for 2.53% of total deaths [3]. A Nationally representative study shows that the prevalence of CKD in the adult population of Nepal was 6% which is associated with several cardiometabolic traits [4]. According to the World Health Organization (WHO) in 2020 kidney disease deaths in Nepal reached 2.67% of the total deaths (i.e.103 in the world) whereas malnutrition causes 0.53% of the total deaths (i.e.59 in the world).

Early detection of disease allows care and management to help prevent morbidity and mortality and improve cost-effectiveness and sustainability. An increased risk of death, mainly from cardiovascular disease, is present in patients with CKD which is not adequately explained by conventional or non-conventional uremia-related risk factors. Undernutrition and nutritional problems among CKD patients have been linked to lower patient survival rates and a lower quality of life. The dietary management of non-dialysis, dialysis, and transplant patients is a particularly difficult area of routine clinical practice, and optimal nutritional status continues to be a poorly understood subject [5]. To achieve optimal nutritional status for patients with chronic kidney disease nutritional support, dietary behaviors and interventions, micronutrients, phosphate control, prevention of malnutrition and protein-energy wasting, and other factors are crucial [6].

Malnutrition often causes weight loss, lowered immunity, and mucosal injury, all of which increase the chance of pathogen invasion. In a vicious cycle, starvation is exacerbated by infection. For instance, the appetite loss typical of acute infectious illness, connected to the production of IL-1 during the acute phase response, further exacerbates anorexia and low caloric intake in patients. Hypoalbuminemia, which is caused by malnutrition, uremia, or the elevated inflammatory response in dialysis patients, may increase susceptibility to infections and ascites in CKD patients. Hypoalbuminemia has also been linked to pneumonia, septicemia, and other inflammatory reactions [7].

A major factor in determining the morbidity and mortality of patients with chronic kidney disease both before and after starting maintenance dialysis therapy is their dietary condition. The evaluation of chronic kidney disease patients’ nutritional status is a crucial component of their care. According to studies, between 18% and 80% of dialysis patients are nutritionally deficient in some way [8]. Numerous clinical practice guidelines for the treatment of individuals with chronic kidney disease have underlined the significance of nutritional status. The techniques and frequency of nutritional evaluation are all covered by American, Canadian, European, and Australian guidelines [9].

Nutritional status and physical activity levels are closely related in an individual. A sedentary lifestyle is encouraged by inadequate food intake, which furthers the vicious cycle of loss of muscular strength and mass, which restricts the quality of life and rehabilitation of CKD patients. Patients with CKD who engage in regular physical exercise and consume enough protein and energy prevent protein-energy waste, promote anabolic effects, and lower associated morbidity and mortality, compared to controls of the same age and sex. CKD patients had lower levels of physical activity and dietary factors [6,10].

There is a paucity of data regarding the assessment of nutritional status and its associated factors among patients with CKD in Nepal. Therefore, this study aims to assess nutritional status and its associated factors among CKD patients in a tertiary care center in Nepal.

## Materials and Methods

### Research design

The study was a descriptive cross-sectional design using primary data. Quantitative method was used in this study. The study was conducted among CKD patients of the Nephrology Department of Tribhuvan University Teaching Hospital. The study population was the patients diagnosed with CKD visiting the nephrology outpatient department of TUTH, Kathmandu. Systematic random sampling was used.

An approximate number of patients visiting Nephrology OPD within one month was generated using the OPD register of the last three years during the same period of data collection (N = 875/month). Inclusion and exclusion criteria were explained. The sampling frame per day was calculated by dividing monthly flow by the total number of OPD running days (875/8 = 109/day). The “k^th^” interval was generated by dividing the total sampling frame of a month by the calculated sample size (875/ 290 = 3.01 ≊ 3). The first sample was chosen by simple random sampling, the lottery method. Other, samples were then taken according to every third interval using a circular systematic random method. 36 samples were chosen from each OPD running day. Since it was very difficult for a researcher to take 36 samples in one day, those patients who were called by doctors again in follow-up OPD and those who were undergoing haemodialysis in TUTH were interviewed on those days rather than on OPD days. Due to this data collection, went on smoothly and hassle-free. Name and hospital numbers of those patients were noted, so as to find those participants on another day. The name and hospital number were then destroyed immediately after data collection. Following all these techniques data collections took place for a month. Data collection began on December 1^st^ 2022 and was completed on December 31^st^, 2022 If the selected sample did not fulfil the inclusion criteria of the study, then consecutive patient fulfilling the inclusion criteria was selected as a sample for the data collection. The sample size was determined using the statistical formula, by Cochran. Sample size (n) = z^2^pq/d^2^ = (1.96)^2^× 0.22 × 0.78 / (0.05)^2^= 263.68 ≊ 264 where, Z = 1.96, considering a 95% confidence level; p = 22% = 0.22 (In a study conducted in Pokhara, Nepal, 22% prevalence of normal nutritional status among CKD patients) [11]; q = 1-p = 78% = 0.78; d = margin of error (5%); Adding a 10% non-response rate, a sample size of 290 was taken.

The inclusion criteria were patients age 18 years and above diagnosed with CKD at least 6 months prior to data collection, since the weight of last 6 months was required for the study. CKD patients with other severe co-morbidities such as heart failure and liver cirrhosis were excluded in order to rule out the determinants of nutritional status due to other major medical reasons. CKD patients in the inpatient department were excluded as they might have obvious limitations for physical activities. Pregnant women were also excluded, so as to prevent them from the possible strain which may occur by the study.

The data was collected using a validated questionnaire: “7 point Subjective Global Assessment tool” [12] for nutritional status and “International Physical Activity Questionaire short form (IPAQ-SF)” [13] for assessing physical activity. Both of these tools are freely available and can be used after acknowledging the authors.

The questionnaire was divided into four sections

1. Socio-demographic characteristics
2. Information on the status of CKD.
3. Information related to assessment of Nutritional Status for CKD.
4. Information related to Physical Activity.

Subjective Global Assessment (SGA) for CKD: It was developed by Detsky et al. [14]SGA is a valid tool for nutritional status and surpasses non-composite nutritional markers, as a predictive indicator for all-cause mortality in both CKD non-dialysis and dialysis patients. It was measured under 7 domains: four using subjective questionnaires and the other three will be recorded with the help of doctors through regular physical examination. The SGA is a tool that quickly and with little technical expertise can determine the nutritional score. The National Kidney Foundation (NKF) Kidney Disease/Dialysis Outcomes and Quality Initiative (K/DOQI) validated and has suggested SGA for use in nutritional assessment in the adult population with CKD due to its advantages [15].

IPAQ-SF: One of the most used self-reported tools for measuring physical activity is the International Physical Activity Questionnaire Short-Form (IPAQ-SF). It comprises seven questions to measure the average amount of time spent each day over the previous seven days standing, walking, and engaging in moderate to strenuous physical activity. The test-retest reliability, concurrent validity, and criterion validity of the IPAQ were created by an international consensus group in 1998 and were examined in 12 different nations [15]. It was also used in the assessment of the physical activity of CKD patients.

Data was collected by the researcher herself using previous records from OPD cards, face-to-face interview techniques and regular physical examinations. In case the patient didn’t carry old cards, a contact number was shared and they were requested to send a photo of the OPD card through mail. The regular physical examinations conducted by the doctors at OPD provided a record of the patient’s physical health. Since, a history and physical examinations are routinely conducted during a doctor’s visit in the OPD of CKD patient, in order to determine whether or not renal replacement therapy might be necessary. Physical examinations were not done merely for research purposes only.

For measuring the weight of the participants Krups mechanical weighing machine was used. Krups is a leading weighing machine manufacturer. The scale shows an accuracy of 0.05 grams and is highly sensitive. It comes with an analogue display. This device is made of mild steel with a powdered coating and brightly coloured mats. Maximum weight capacity of 180 kilograms. Calibration of the instrument was done after each measurement. It is WHO certified and also ISO-9000-2008 certified.

### Data Processing and Analysis

Data collection was done by the researcher herself. The researcher was responsible for the completeness and correctness of the collected data. Data compilation, checking, editing and coding were carried out following the data collection. Data were systematically coded and entered into IBM SPSS Version 25 where consistency was checked and cleaning and editing of data were done. All analysis was performed using IBM SPSS version 25.

Descriptive analysis: Frequency, percentage, mean and standard deviation, were used to assess nutritional status and to find out the level of physical activity among CKD patients.

Inferential statistics: The chi-square test was used to assess the association between dependent and independent variables. Cross-tabulation was used for evaluating the relationship between a dependent variable and other explanatory variables. Odds ratio at 95% confidence level and Chi-square with p-value < 0.05 was taken.

Logistic regression was applied to determine the association between dependent and independent variables. The adjustment of variables which might be probable confounders to the dependent variable was done with binary logistic regression using the entry technique. Variables that were found significant at a 5% level of significance in bivariate analyses were entered into the multivariate analysis based on the Hosmer-Lemeshow goodness of fit test for logistic regression criteria. The Variance inflation factor (VIF) and Tolerance were examined using collinearity diagnostics before variables were added to the regression model. No difficulties of collinearity among the explanatory variables were detected.

### Validity

The data was collected, using a validated and standard questionnaire, the Subjective Global Assessment tool in order to assess nutritional status. The study instrument was translated from English to Nepali. Translational validity was assured using forward and backward translation techniques with the help of two experts (A language expert and a Subject expert). A language expert having done a Master’s in the Nepali language with equal proficiency in English was consulted. For the subject expert doctors of the Nephrology department were consulted. Face validity was assured by pretesting among 30 participants with CKD, and necessary modifications were done.

### Reliability

The reliability of the SGA was established by pre-testing the tool in 30 participants with CKD of TUTH. Hospital Identification number and name of pretested samples were taken, so that they could be excluded during data collection. This information was later destroyed after the completion of data collection. During pretesting, the translation and understanding of the questionnaire were checked, and corrections were made to the wording of the questions. Pretesting helped to access the adequacy of the questionnaire and to identify logistical challenges that might have occurred during the actual study. Internal consistency was ascertained by Cronbach’s alpha. The Cronbach alpha value of 0.8 was obtained. The Cronbach value of the original scale is 0.86.

### Bias

Recall bias could have occurred as participants have to remember information such as their dietary intake, GI symptoms, and functional status of the past 2 weeks. They also need to remember the physical activity done by them in a week. To minimize this during the study, the participants were given enough time to remember their history and physical activity performed by them. Also, the participants were allowed to ask and cross-check with their other family members or visitors, so that appropriate information about them could be collected.

### Consent

The principal investigator was responsible for obtaining informed consent from participants. Assurance was provided to the participants that the information was kept strictly confidential, their names won’t be used anywhere and the information obtained would be used for academic purposes only. Participants were explained that the participation would be completely voluntary and there was no any compulsion and they were free to withdraw from the research at any time. The participants would not be exposed to any risks in this research. They were only be requested to provide honest responses. After agreeing on these given points, the interview began.

### Ethical Consideration

Ethical consideration was taken from the Institutional Review Committee, Institute of Medicine [Ref: 254(6-11)E2]. Permission letter was taken from the Nephrology department and hospital administration of TUTH. Written informed consent was taken from the participants. In order to ensure privacy participants were asked separately and the provided data won’t be shared to anyone. It was limited to researchers only. For the confidentiality of participants, each instrument was coded randomly and aggregate reporting was done. Malnourished patients after consultation with doctors were referred to a dietitian for further management. Also, the tools used for data collection could be used freely with acknowledgement.

## Results

A total of 290 patients were included in the study. Table 1 provides a description of the socio-demographic characteristic of the study population. Nearly half (49.7%) of the participants were between ages 41 to 65 years and 125 participants (43.1%) were of the age group 18-40 years. The mean age of participants was 43.92 ± 14.87 years. The majority of the participants (65.2%) were males in this study. More than half (58.6%) had education up to the secondary level and higher whereas 41.4% had education below the secondary level. Most of the patients (74.8%) were employed in various sectors like service, business, agriculture and manual workers, whereas one forth (25.2%) participants were unemployed. One third (33.1%) were from privileged ethnic groups such as Brahmin and Chhetris, on the other hand remaining (66.9%) were underprivileged groups such as Janajati, Madhesi, Dalit, and Muslims. The majority of the participants (80.3%) were Hindus and the remaining (19.7%) were non-Hindus such as Buddhist, Muslim, Christian and Kirat.

**Table 1.**
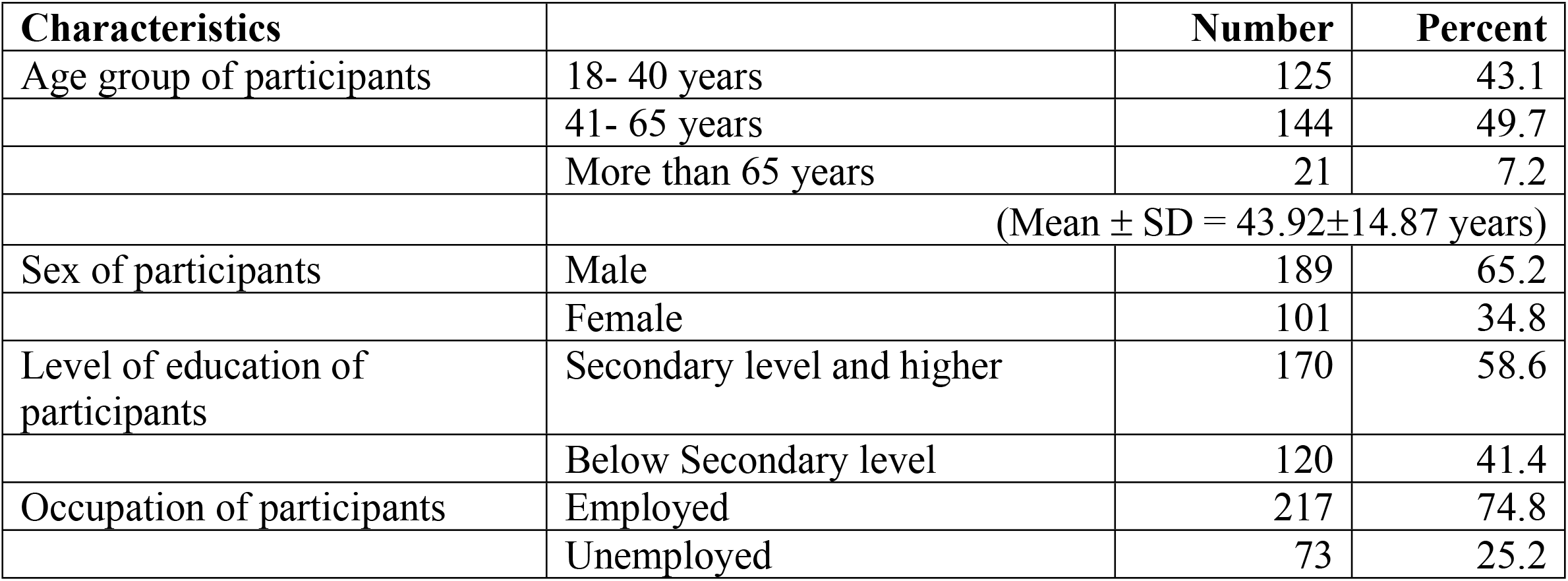
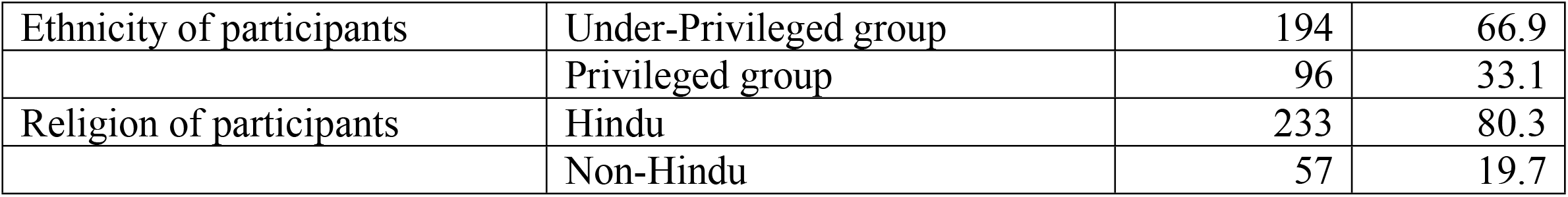
Socio-demographic characteristics of the study population.

Among the 290 participants, 244 (84.1%) had late-stage CKD. Nearly, half of the participants (49.3%) were diagnosed before or equal to two years from the day of data collection. The majority of the participants (59%) were on hemodialysis. Out of 171 participants on hemodialysis, greater than two-third (70.17%) had been doing it for less than two years and the remaining (29.83%) had been doing hemodialysis for more than and equal to two years. Nutritional status of the patients was assessed using seven components – weight loss over six-month, dietary intake, gastrointestinal symptoms, functional status, muscle storage, fat storage and edema. Amongst the 290 participants, one-fourth of the participants (26.9%) were malnourished.

The level of physical activity was assessed using IPAQ-SF and the subjective data were interpreted using MET per week. More than half of the participants (53.4%) had a moderate level of activity. Only one-fourth of the participants (25.2%) had high activity levels while 21.4% had low activity levels.

Table 2 presents the association of socio-demographic factors with the malnutrition of CKD patients. The factor that showed statistical association with the malnutrition of the CKD patients includes the sex of the participants (p-value .006, 95% CI 1.225-3.554). The socio-demographic factors such as age, level of education, occupation, ethnicity and religion of the participants did not show any significant association with the malnutrition of the CKD patients.

**Table 2.**
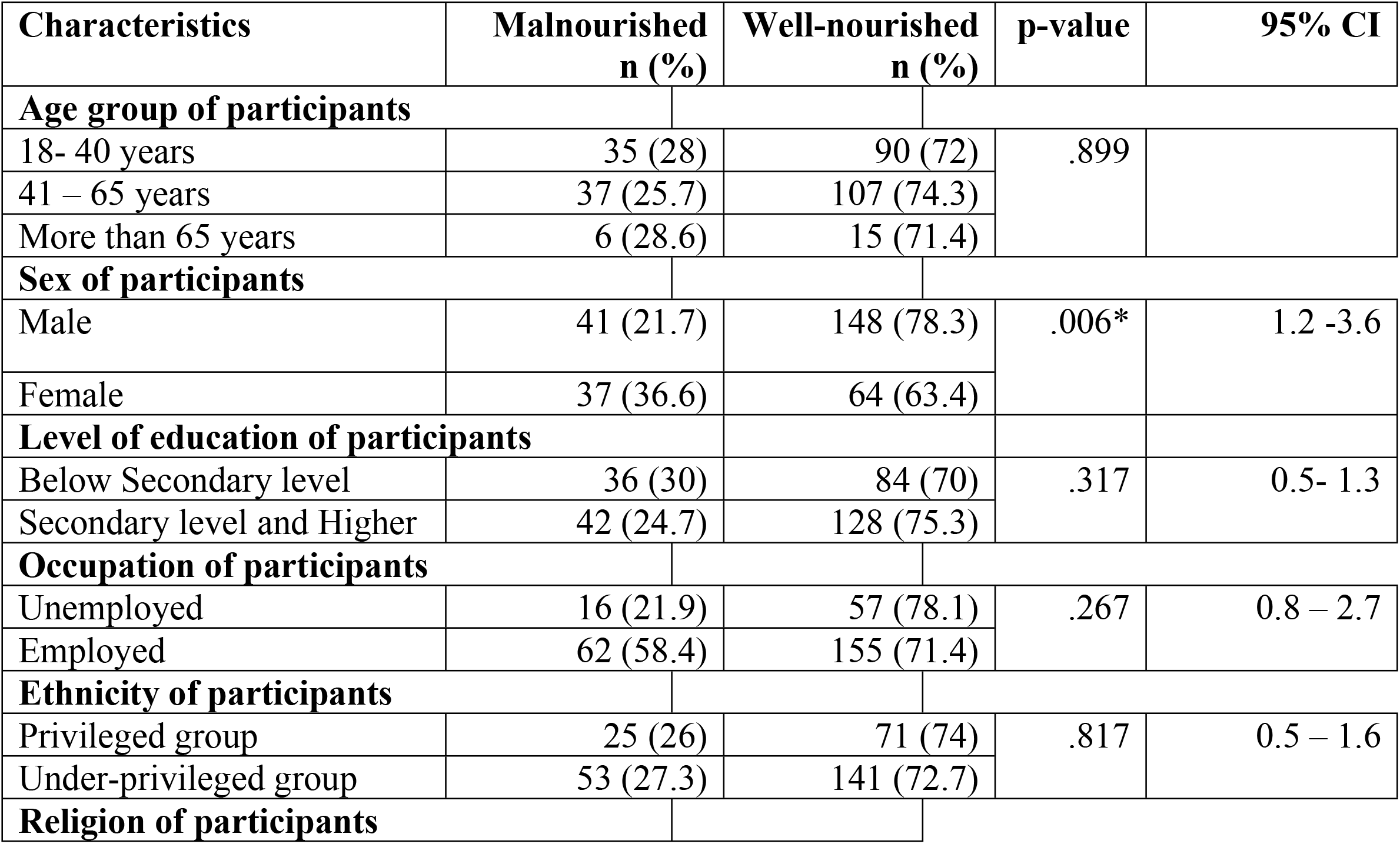
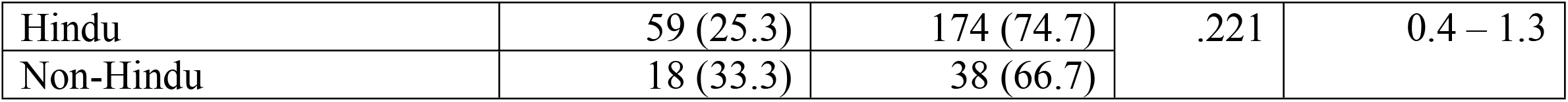
Association of socio-demographic factors with malnutrition.

Table 3 describes the association of CKD status with the malnutrition of CKD patients. In the cross-tabulation of various characteristics of CKD status, a statistically significant association was found with the duration of hemodialysis among the participants undergoing hemodialysis (p-value.002, 95% CI 0.98 – 0.628). Other characteristics of the CKD status such as stages of CKD, period of CKD diagnosis and status of hemodialysis were not significantly associated with the malnutrition of CKD patients.

**Table 3.**
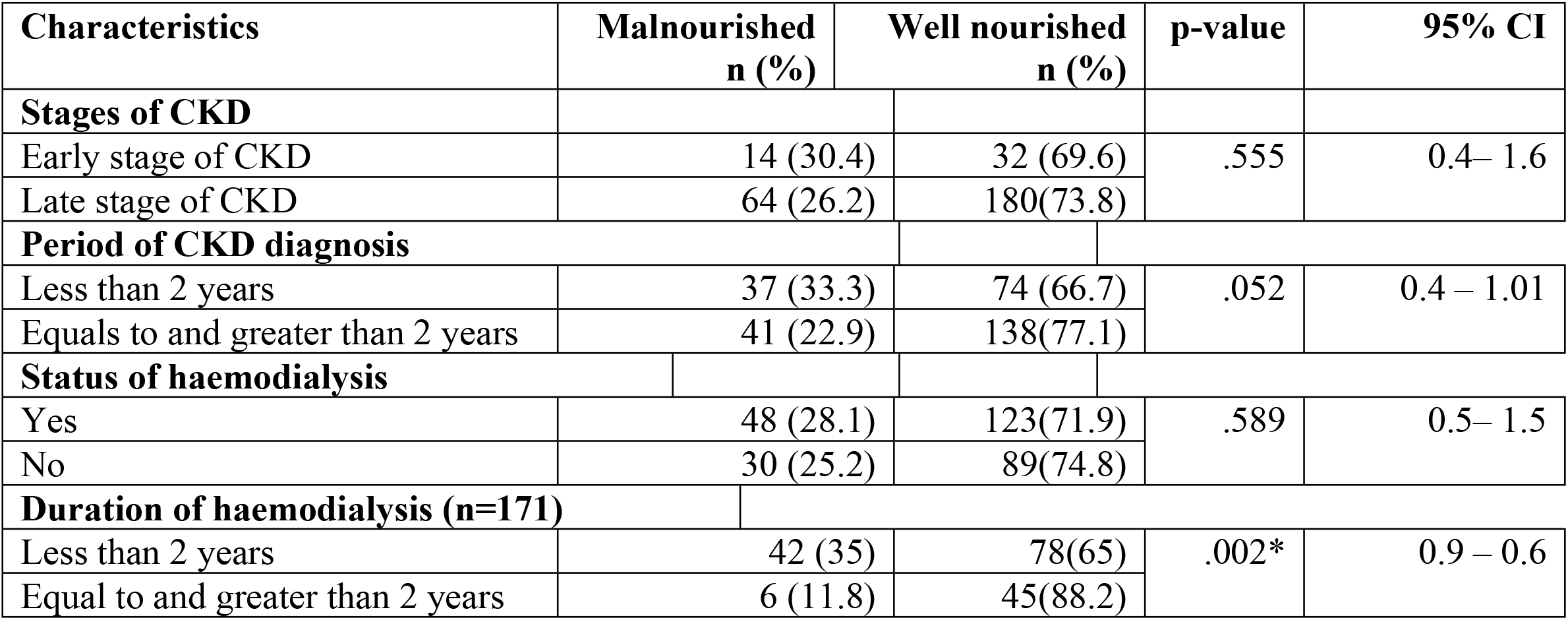
Association of CKD status with malnutrition.

Table 4 shows the association of physical activity with the malnutrition of CKD patients. Physical activity was significantly associated with the malnutrition of the CKD patients (p-value <.001).

**Table 4.**
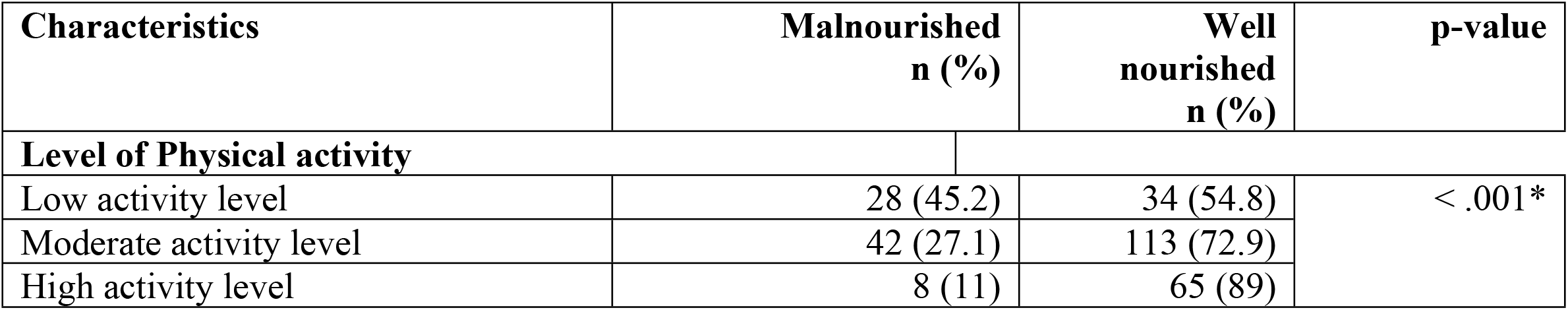
Association of physical activity with malnutrition.

The adjusted relationship of the explanatory variables with malnutrition in CKD patients were assessed at 95% CI in the multivariate analysis. The adjustment of factors which could be possible confounders to the dependent variable was done with binary logistic regression using enter method. Three variables that were found significant at a 5% level of significance in bivariate analyses were entered into multivariate analysis. One variable was taken to binary logistic regression, based on the criteria (p–value<0.1) given by the Hosmer-Lemeshow goodness of fit test for logistic regression. Prior to the inclusion of variables in the regression model, the Variance inflation factor (VIF) and Tolerance were checked using collinearity diagnostics. No problems of collinearity among the explanatory variables were identified (the highest observed VIF was 1.237 and the lowest observed Tolerance was 0.808).

Table 5 shows the adjusted ORs, crude ORs, their 95% CI, and p-value of the factors associated with malnutrition in CKD patients with and without hemodialysis. Results from regression analysis showed that the sex of participants and physical activity were significant with malnutrition in CKD, whereas the period of diagnosis of CKD was taken to binary logistic regression, based on the criteria (p–value<0.1) given by Hosmer-Lemeshow goodness of fit test for logistic regression. Male participants were less likely to have malnutrition compared to female participants with CKD (AOR 0.451, p-value0.005). Likewise, the period of diagnosis of CKD was not particularly useful in predicting malnutrition (p-value 0.071 >0.05). The results illustrate that participants having low activity levels were about 7times more likely to have malnutrition compared to those having high activity levels (AOR 7.022, p-value <0.000). Similarly, participants having moderate activity levels were about 3 times more likely to have malnutrition compared to those having high activity levels (AOR 3.263, p-value 0.005). Based on the estimates from multivariate analysis, the final regression model was obtained as:

**Table 5.**
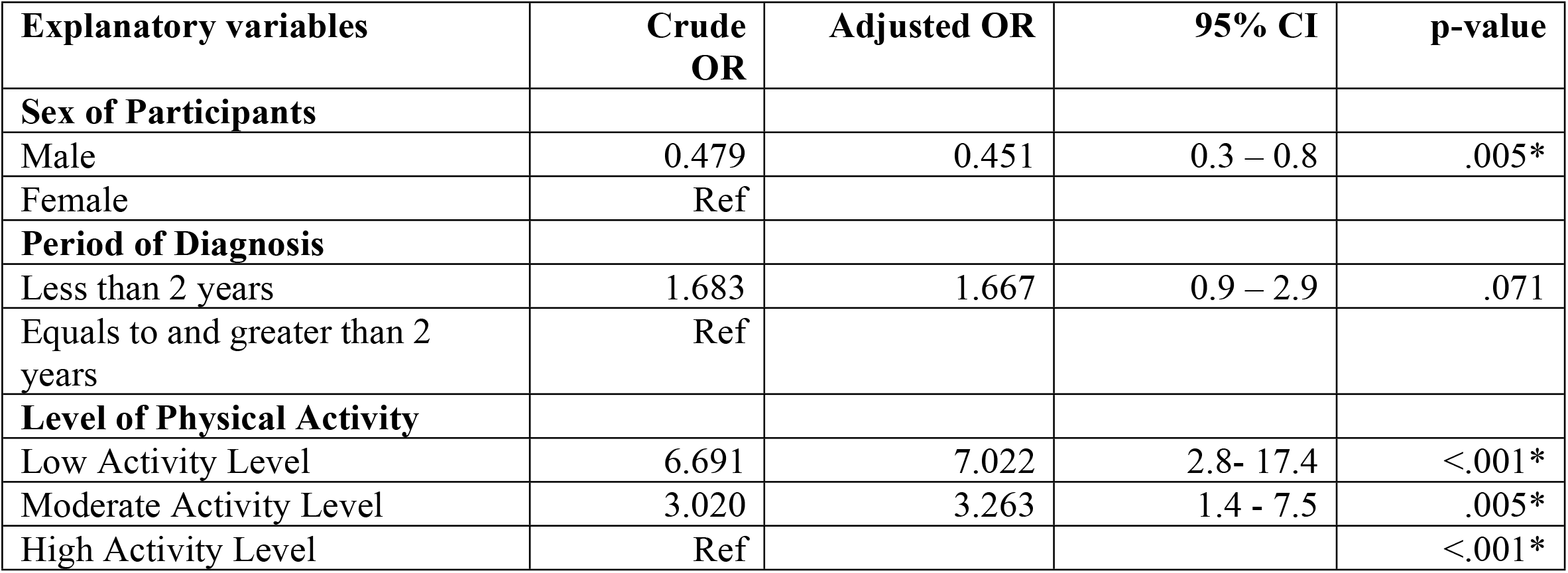
Adjusted relationship of the explanatory variables with malnutrition in the CKD patients (with and without haemodialysis)

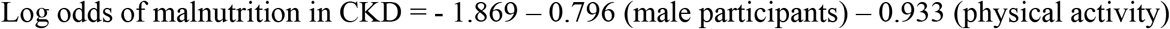

Hosmer and Lemeshow’s Chi-square statistic in the model showed no significant difference between the observed and predicted probabilities (p-value 0.157). This indicated a good model fit. Also in the model, a Nagelkerke (pseudo) R Square value of 0.151 was observed which indicates that 15.1% of the variability in the malnutrition of the CKD patients (with or without hemodialysis) was explained by the explanatory variables.

Table 6 describes the adjusted relationship of the explanatory variables with malnutrition in CKD patients undergoing hemodialysis. The results depict that participants who were under hemodialysis for less than 2 years were about 2.8 times more likely to have malnutrition compared to those under hemodialysis for greater than and equal to 2 years (AOR 2.826, p-value 0.044).

**Table 6.**
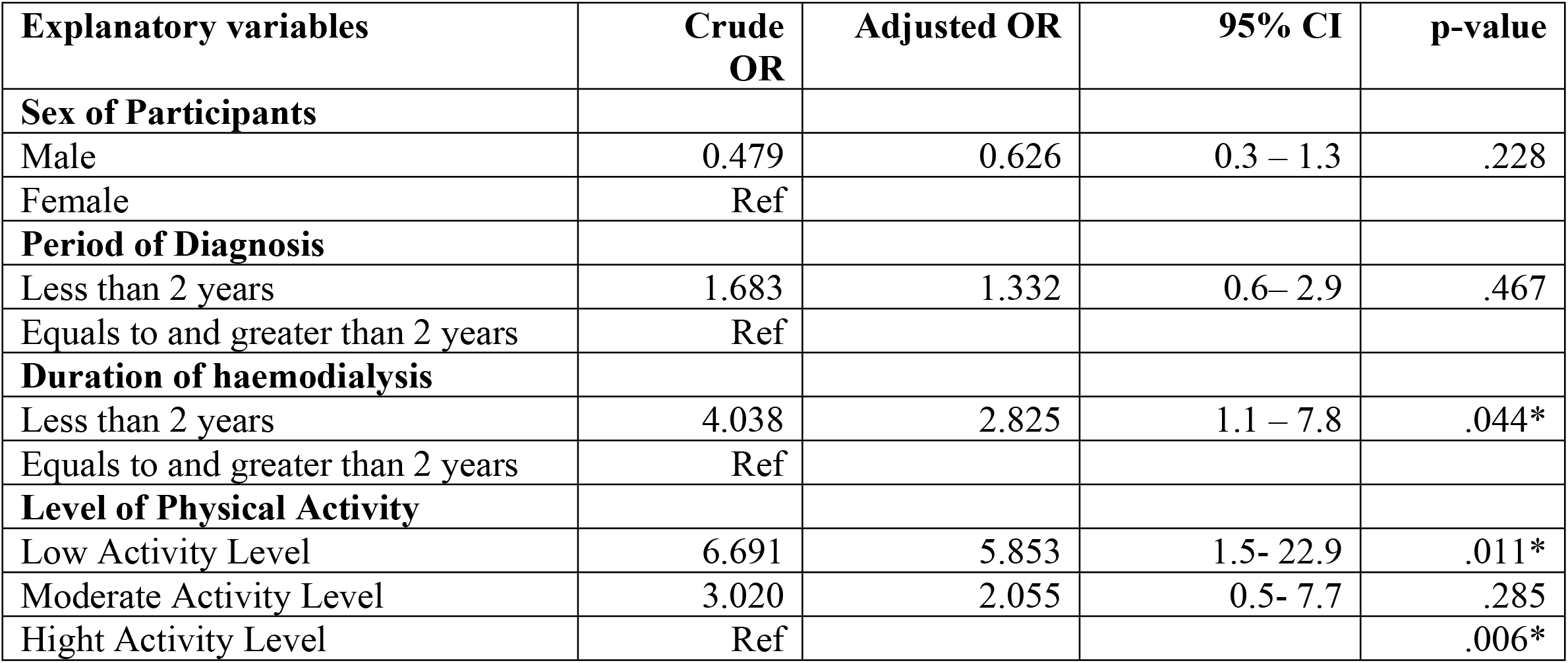
Adjusted relationship of the explanatory variables with malnutrition in the CKD patients undergoing haemodialysis.

Furthermore, participants having low activity levels were about 5.8 times more likely to get malnutrition compared to those having high activity levels (AOR 5.853, p-value 0.011). The sex of participants and period of diagnosis of CKD was not particularly useful in predicting malnutrition in CKD patients undergoing hemodialysis (p-value > 0.05).

Based on the estimates from multivariate analysis, the final regression model was obtained as:

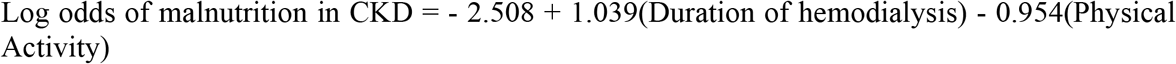

Hosmer and Lemeshow’s Chi-square statistic in the model showed no significant difference between the observed and predicted probabilities (p-value 0.131). This indicated a good model fit. Also in the model, Nagelkerke (pseudo) R Square value of 0.189 was observed which indicates that 18.9% of the variability in the malnutrition of the CKD patients under hemodialysis was explained by the explanatory variables.

## Discussion

This study was conducted to assess the factors associated with malnutrition in CKD patients attending the outpatient department of Tribhuvan University Teaching Hospital. The factors associated with malnutrition in CKD patients as identified in this study includes sex of participants and the level of physical activity. Similarly, the factors associated with malnutrition in patients under hemodialysis include duration of hemodialysis and level of physical activity.

This study showed, 26.9% of participants with CKD had malnutrition which is similar to the various studies which state, the prevalence of malnutrition has been estimated to range from 20 to 75% depending on the diagnostic criteria and the participants [16]. Malnutrition is more common in the late stages of CKD, with a prevalence of 23 to 75% [17]. Researchers came to the conclusion that CKD patients have a high prevalence of malnutrition. A systematic review and meta-analysis conducted across 61 countries showed the global prevalence of malnutrition in CKD patients to be 42.7% while the prevalence of malnutrition in India among CKD patients was found to be 56.7% which is more than our study [18]. Numerous studies have shown that malnourished CKD patients have higher death and morbidity rates. It is noteworthy to observe that malnutrition is rarely listed as a cause of death in CKD patients. However, a growing body of research indicates that these individuals’ dietary state has a significant impact on their prognosis [16].

In this study, 21.4% of the participants with CKD had low activity level which almost resembles the observational multi-center study conducted in the United Kingdom where the prevalence of insufficient physical activity was 6% −34% and worsened with disease progression i.e., patients in stage 1-2 were more active than those in stage 4-5 [19].

Evidence from this study indicates male participants were less likely to have malnutrition compared to female participants with CKD which is a contrast to the meta-analysis which signifies that compared to females, men were more commonly affected by malnutrition [18]. Also, another study from India showed an insignificant difference between malnutrition in CKD and the sex of the participants [20]. The sex of participants was not particularly useful in predicting malnutrition in CKD patients undergoing hemodialysis in Iran which is identical to this study [21]. This may be related to the included research participants and the malnutrition evaluation standards or may have been related to gender bias related to food in Nepal.

In this study duration of hemodialysis is associated with malnutrition, the lesser the duration (< 2 years) of hemodialysis more is the likelihood of having malnutrition. The intake of proteins, fats, and the majority of nutrients, including minerals and vitamins, was lower at the beginning of hemodialysis. Studies found that CKD patients exhibited protein-energy malnutrition, during the early phase of dialysis [22]. Undernutrition results from poor intake due to anorexia, nausea, and vomiting; it typically gets better with sufficient dialysis and suitable nutritional support. Body mass index grew slowly, peaked during the tenth and twelfth years of dialysis, and then decelerated after 16 years of hemodialysis [23]. The observed association may have been due to a greater number of participants in pre-dialysis and early phases of hemodialysis. In opposition to this, a study in Tanzania showed those patients on longer than 4 years of hemodialysis have a higher chance of malnutrition [22].

Various study confirms the positive relationship between malnutrition and low level of physical activity in CKD patients which resembles this study. Clinically, physical inactivity is a long-standing issue for CKD patients, especially for those receiving dialysis. Physical activity boosts the anabolic effects of nutritional treatments. Increases in physical performance and activity may help improve patients’ nutritional health [24]. Better physical activity was associated with better nutritional status and lower comorbidity in patients undergoing hemodialysis [25]. Inactivity is a main cause of muscle atrophy and may be a factor in the CKD population for muscular abnormalities and decreased functional status [26].

Several predictive factors that were significantly associated with malnutrition in the previous study were not significant in our studies such as age, level of education, occupation, ethnicity and religion. For example, a study from Nigeria showed malnutrition significantly increases with an increase in age, malnutrition was higher in the elderly compared to younger and middle-aged patients [27]. Higher prevalence was significantly associated with unemployment and education [27]. The research population size, recruited age, data collection techniques, socioeconomic status,patients’ food habits, and medical intervention may all differ from those of the aforementioned variables and those of the current study, which could account for the variations between the two.

All of the results previously presented showed how important it is for multidisciplinary healthcare professionals to work together to treat CKD patients’ risk of malnutrition. Frequent nutritional screening, routine patient education and anthropometric monitoring during each follow-up should be performed with appropriate documentation by the health care professionals.

To the best of our knowledge, this is one of the first few studies attempting to identify the factors associated with malnutrition among CKD patients in Nepal. In this study, there could be some chances of recall bias since the self-reported method was used and the recall period for some information. Although the findings of this study were consistent with a large body of literature from various low and middle-income countries, we couldn’t relate these findings to our national context owing to the limited number of literature available for Nepal. Despite these limitations, the results of the study will add to the knowledge base on malnutrition in CKD patients and generally be useful to policymakers, decision-makers and those in academia.

## Conclusion

This study estimated the nutritional status and status of physical activity and also determined factors associated with nutritional status among patients with chronic kidney disease. Malnutrition among CKD patients was between 21.8% to 32% and the level of physical activity was low among 16.8% to 26.1% of participants. Female participants with low activity levels and those undergoing hemodialysis for less than two years are more likely to have malnutrition. It is thus recommended that patients with CKD are advised to engage in at least mild to moderate levels of physical exercise. Every six months, health professionals are advised to do nutritional assessments and supply them with diet charts if needed. It is also recommended to assess and treat malnutrition in the non-dialysis population. Caretakers are advised to treat males and females equally because both are equally vulnerable to malnutrition.

## Data Availability

All relevant data are within the manuscript and its supporting information files..

## Acknowledgments

We would like to express our deep gratitude to Prof. Dr. Amod Kumar Poudyal, Prof Dr Mahesh Raj Sigdel and Dr Midhan Shrestha for their valuable support and suggestions. Further, we would like to express our sincere thanks to Nephrology Department, Tribhuvan University Teaching Hospital for providing us permission for data collection, and hospital administration for coordinating with staff members for data collection at the Out Patient Department of Nephrology.

